## Supplemental figures and tables for "Contrasting factors associated with COVID-19-related ICU admission and death outcomes in hospitalised patients by means of Shapley values"

### Supplemental tables and figures

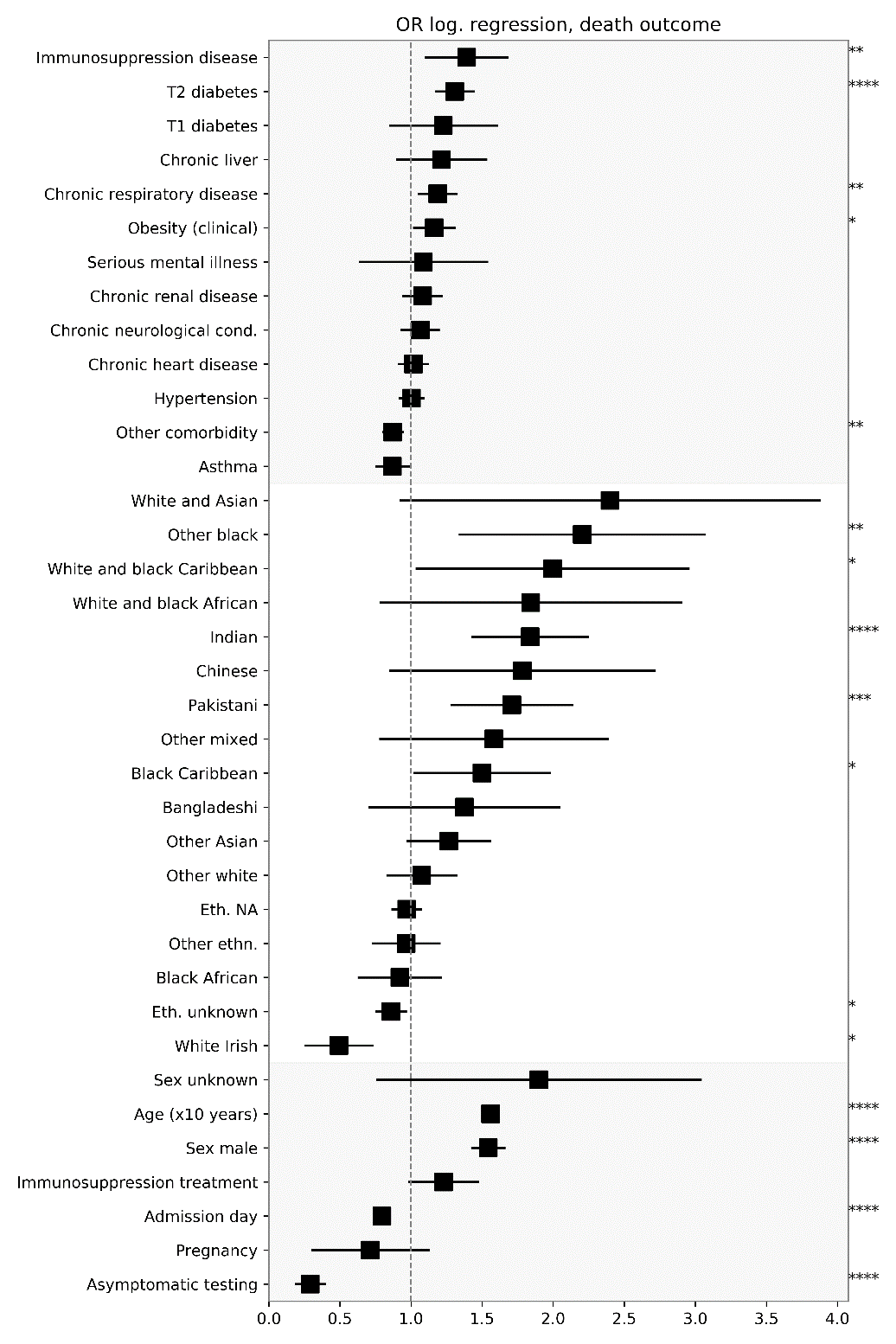

Figure S1. Odd ratios (ORs) for death outcomes from Table 3. Features are grouped in comorbidities, ethnicities, and others (top to bottom). Significance star codes are `*` P ≤ 0.05, `**` P ≤ 0.01, `***` P ≤ 0.001, `****` P ≤ 0.0001.

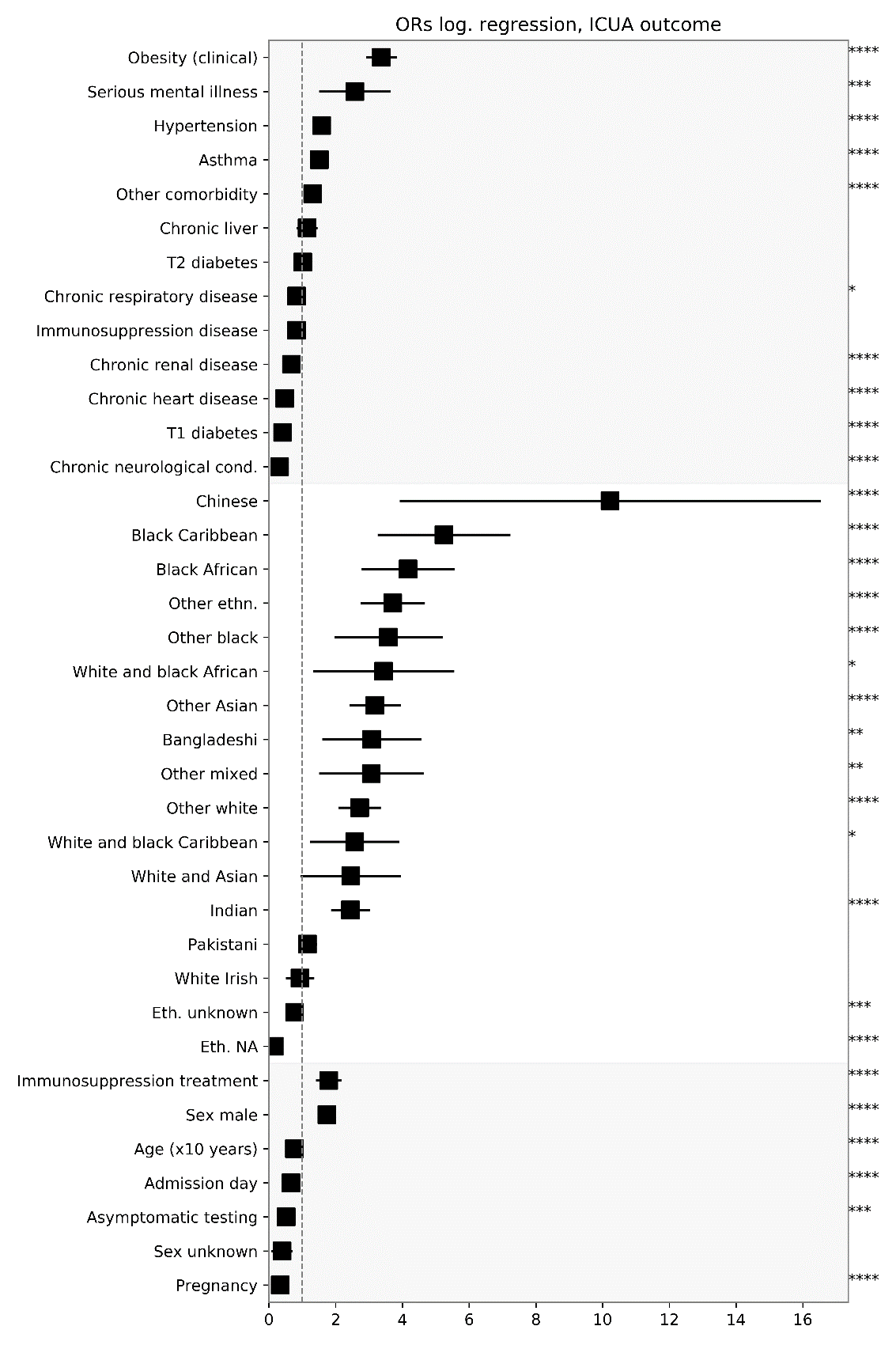

Figure S2. Odd ratios (ORs) for ICUA outcome from Table 3. Features are grouped in comorbidities, ethnicities, and others (top to bottom). Stars are codes for significance (* P ≤ 0.05, **P ≤ 0.01, ***P ≤ 0.001,**** P ≤ 0.0001).

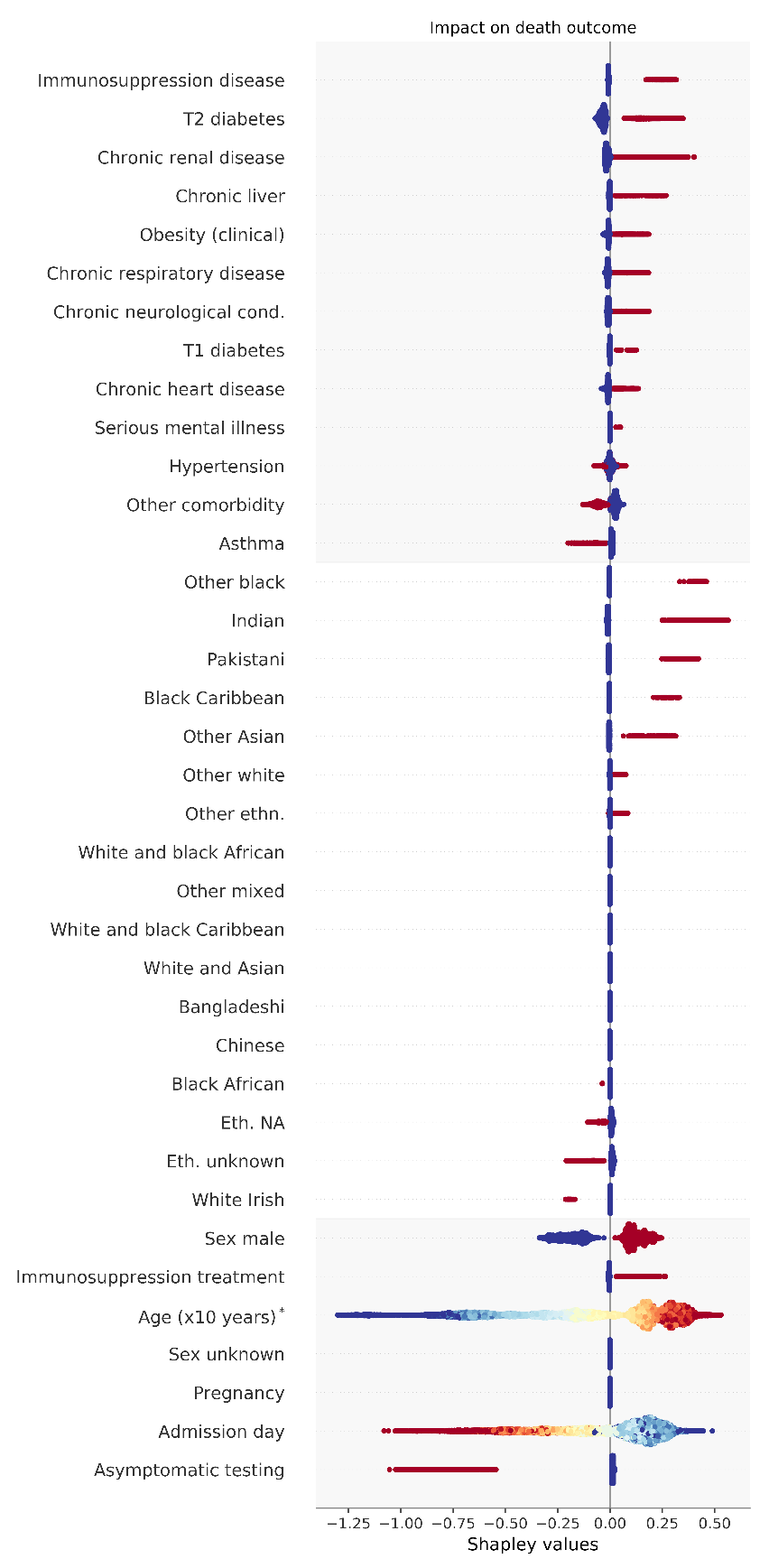
Figure S3. Summary plot of Shapley values for impact on death outcome. For each potential risk factor, the Shapley values of each patient are represented as a swarm plot distribution. Colors from red to blue indicate the value of the underlying variable (in binary variables, red color means feature is present, blue otherwise; in age feature, red to blue shades correspond to old to young ages; in admission day, red to blue shades correspond to early to late dates). Features are grouped in comorbidities, ethnicities, and others. Within each group, the factors are ranked according to the importance score *Imp* (see Table 3); in other words, upper in the list are the conditions most likely to be associated with the worst outcome. Immunosuppression by disease, type-2 diabetes mellitus, being male, and chronic liver, renal, respiratory and neurological conditions consistently appear to have positive impact to death outcome for all patients. Asthma was found to have negative impact on death for all patients. *Shapley values for age (x10 years) are scaled by a factor 0.5 to fit the plot range.

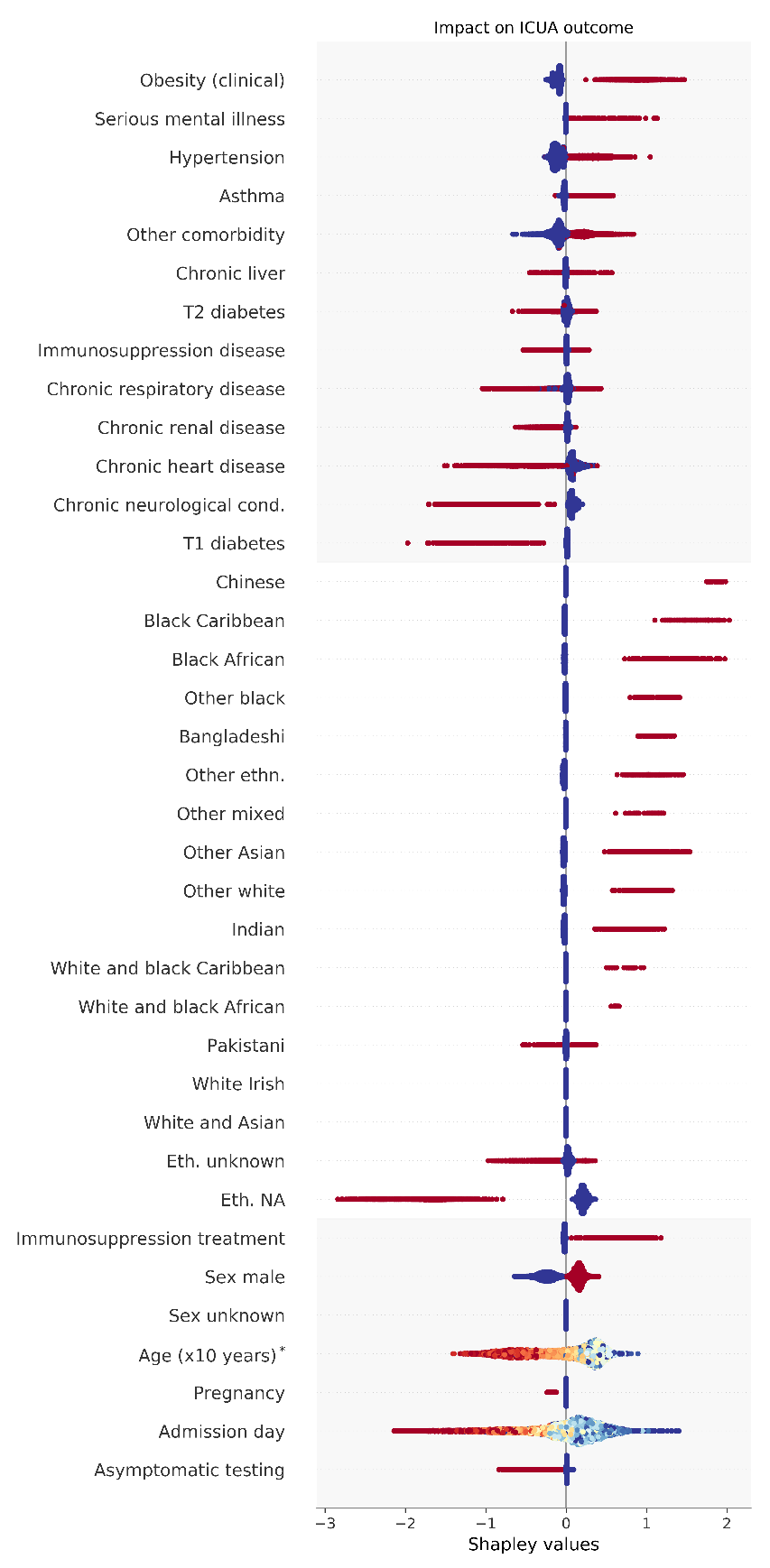

Figure S4. Summary plot of Shapley values for impact on ICUA outcome. Colors and keys as in Figure S3. The presence of obesity, hypertension, immunosuppression treatment, and “other comorbidity” were clear indicators of ICUA outcome for all patients. *Shapley values for age (x10 years) are scaled according to Figure S3.

Table S1. Estimated odd ratios (ORs) and variance inflation factors (VIFs) from logistic regression, importance (Imp) scores of death, and Benjamini-Hochberg (BH) significance test results adjusted for intensive-care unit admission (ICUA).

|  | **OR** | **2.50%** | **97.50%** | **Pr(>\|z\|)** | **VIF** | **Imp** | **BH test** |
| --- | --- | --- | --- | --- | --- | --- | --- |
| ICUA | 2.25 | 2.04 | 2.48 | 0.00 | 1.48 | 0.39 | TRUE |
| **Comorbidities:** |  |  |  |  |  |  |  |
| Immunosuppr. disease | 1.45 | 1.14 | 1.84 | 0.00 | 1.05 | 0.22 | TRUE |
| T1 diabetes | 1.44 | 0.99 | 2.08 | 0.05 | 1.02 | 0.12 | FALSE |
| T2 diabetes | 1.32 | 1.18 | 1.48 | 0.00 | 1.19 | 0.16 | TRUE |
| Chronic neurological cond. | 1.24 | 1.08 | 1.43 | 0.00 | 1.10 | 0.15 | TRUE |
| Chronic liver | 1.24 | 0.91 | 1.67 | 0.17 | 1.02 | 0.21 | FALSE |
| Chronic respiratory disease | 1.23 | 1.08 | 1.40 | 0.00 | 1.10 | 0.12 | TRUE |
| Chronic renal disease | 1.13 | 0.98 | 1.31 | 0.09 | 1.15 | 0.15 | FALSE |
| Chronic heart disease | 1.11 | 0.99 | 1.25 | 0.06 | 1.26 | 0.05 | FALSE |
| Serious mental illness | 1.03 | 0.60 | 1.71 | 0.92 | 1.02 | 0.00 | FALSE |
| Obesity (clinical) | 0.99 | 0.87 | 1.14 | 0.94 | 1.15 | 0.00 | FALSE |
| Hypertension | 0.94 | 0.85 | 1.03 | 0.19 | 1.27 | -0.03 | FALSE |
| Other comorbidity | 0.84 | 0.77 | 0.92 | 0.00 | 1.17 | -0.08 | TRUE |
| Asthma | 0.82 | 0.71 | 0.96 | 0.01 | 1.05 | -0.16 | TRUE |
| **Ethnicities:** |  |  |  |  |  |  |  |
| White and Asian | 2.08 | 0.79 | 5.44 | 0.13 | 1.01 | 0.15 | FALSE |
| Other black | 1.84 | 1.11 | 3.05 | 0.02 | 1.02 | 0.26 | TRUE |
| White and black Caribbean | 1.72 | 0.89 | 3.31 | 0.10 | 1.01 | 0.12 | FALSE |
| Pakistani | 1.69 | 1.26 | 2.27 | 0.00 | 1.04 | 0.33 | TRUE |
| White and black African | 1.59 | 0.66 | 3.62 | 0.28 | 1.01 | 0.11 | FALSE |
| Indian | 1.59 | 1.22 | 2.06 | 0.00 | 1.05 | 0.29 | TRUE |
| Other mixed | 1.36 | 0.66 | 2.66 | 0.38 | 1.01 | 0.04 | FALSE |
| Chinese | 1.31 | 0.61 | 2.76 | 0.48 | 1.01 | 0.00 | FALSE |
| Eth. NA | 1.23 | 1.09 | 1.39 | 0.00 | 1.19 | 0.09 | TRUE |
| Black Caribbean | 1.19 | 0.80 | 1.74 | 0.39 | 1.02 | 0.04 | FALSE |
| Bangladeshi | 1.18 | 0.60 | 2.23 | 0.63 | 1.01 | 0.05 | FALSE |
| Other Asian | 1.05 | 0.80 | 1.37 | 0.74 | 1.05 | 0.04 | FALSE |
| Other white | 0.92 | 0.70 | 1.20 | 0.54 | 1.03 | 0.01 | FALSE |
| Eth. unknown | 0.90 | 0.78 | 1.04 | 0.15 | 1.07 | -0.07 | FALSE |
| Other ethn. | 0.81 | 0.60 | 1.07 | 0.15 | 1.04 | -0.15 | FALSE |
| Black African | 0.74 | 0.50 | 1.07 | 0.12 | 1.03 | -0.18 | FALSE |
| White Irish | 0.51 | 0.26 | 0.95 | 0.04 | 1.00 | -0.39 | FALSE |
| **Other:** |  |  |  |  |  |  |  |
| Sex unknown | 2.09 | 0.82 | 5.27 | 0.12 | 1.01 | 0.39 | FALSE |
| Age (x10 years) | 1.67 | 1.62 | 1.73 | 0.00 | 1.44 | 0.01 | TRUE |
| Sex male | 1.45 | 1.34 | 1.58 | 0.00 | 1.05 | 0.11 | TRUE |
| Immunosuppr. treatment | 1.14 | 0.91 | 1.43 | 0.24 | 1.06 | 0.09 | FALSE |
| Admission day | 0.84 | 0.80 | 0.87 | 0.00 | 1.09 | -0.20 | TRUE |
| Pregnancy | 0.83 | 0.34 | 1.81 | 0.66 | 1.01 | 0.00 | FALSE |
| Asymptomatic testing | 0.31 | 0.19 | 0.47 | 0.00 | 1.04 | -0.73 | TRUE |

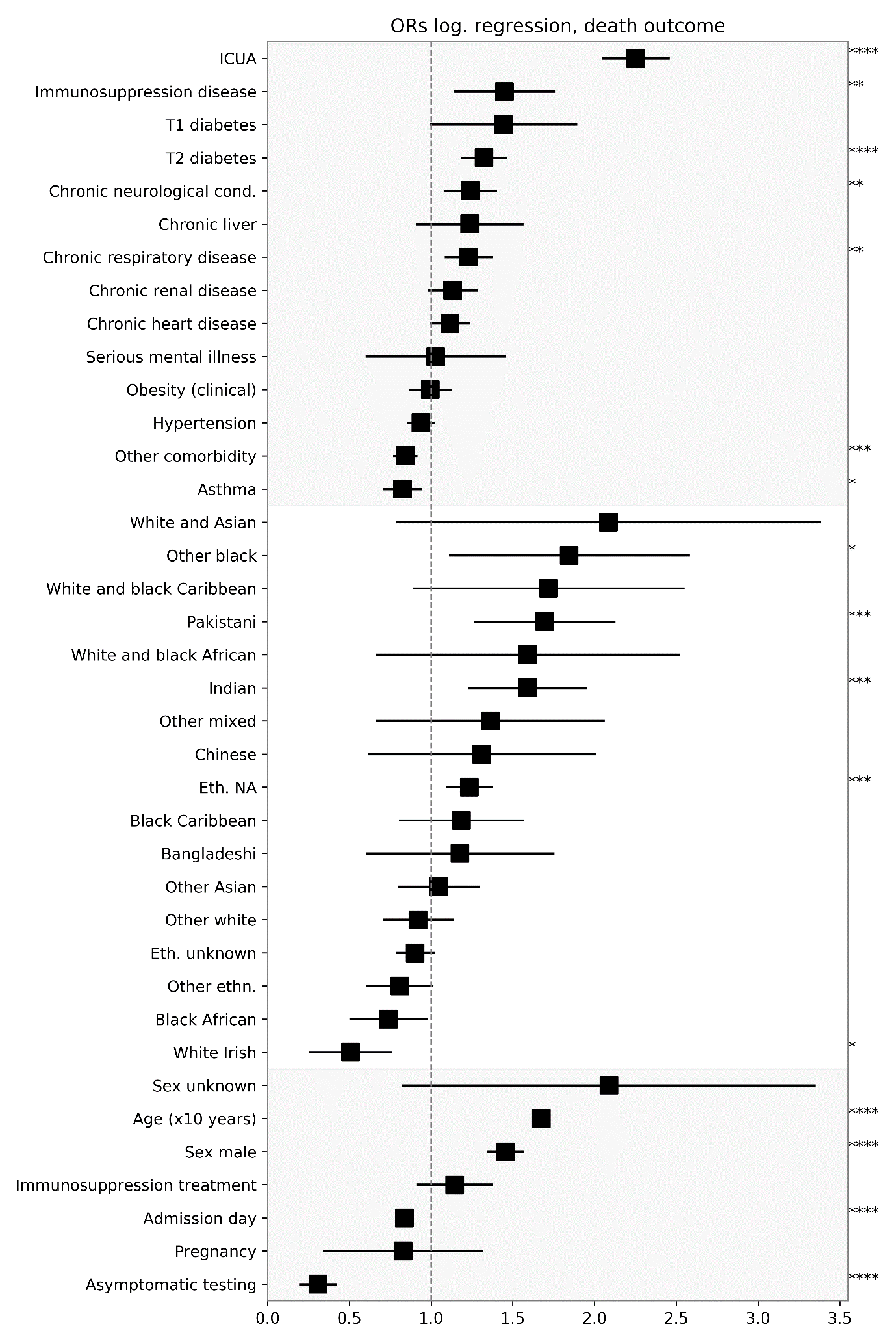

Figure S5. Odd ratios (ORs) for death outcome from Table S1. Features grouping and stars codes for significance as in Figures S1 an S2.

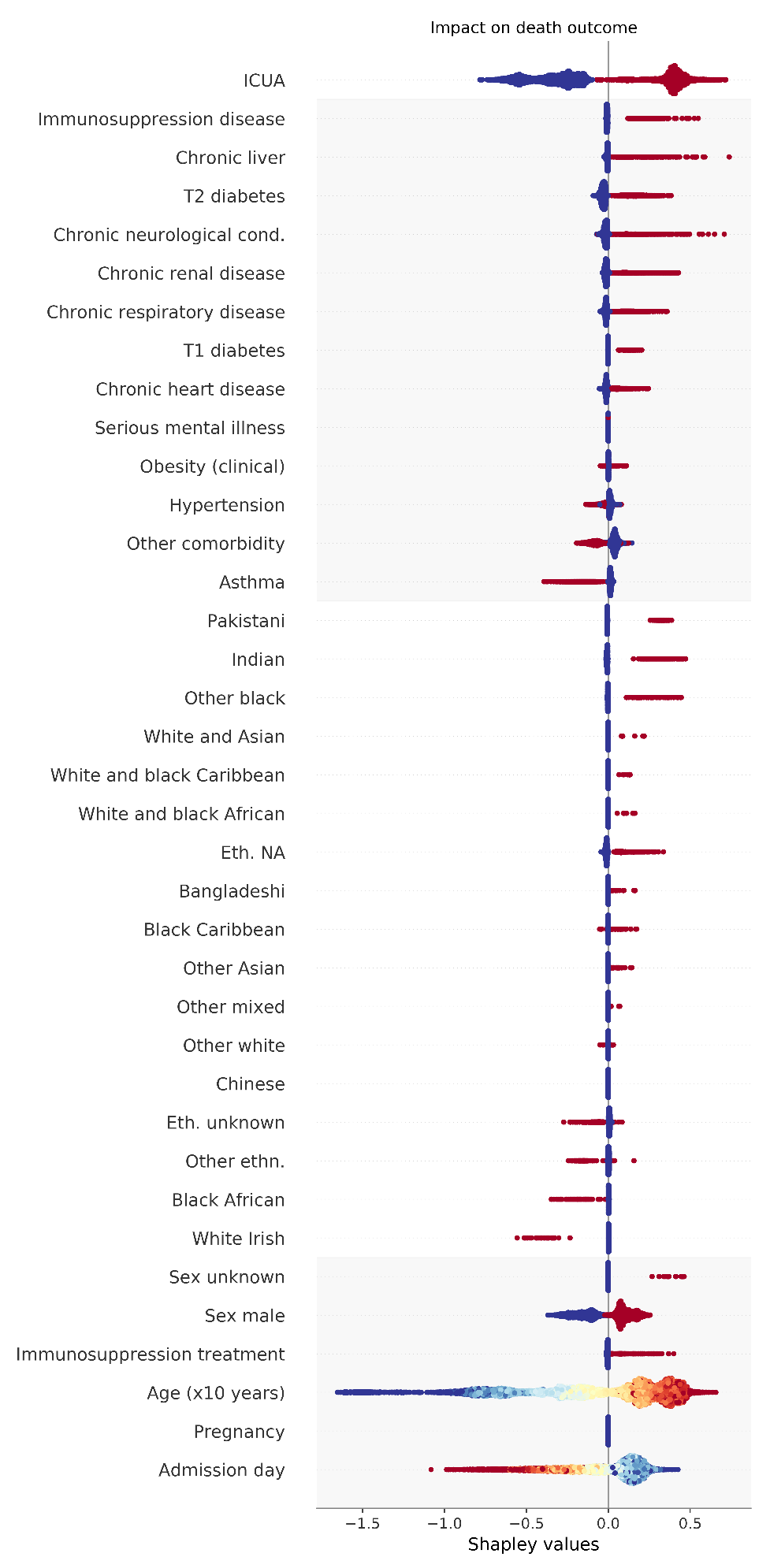

Figure S6. Summary plot of Shapley values for impact on death outcome stratifying on ICUA. Colors and keys as in Figures S3 and S4.
